# Meteorological factors correlate with transmission of 2019-nCoV: Proof of incidence of novel coronavirus pneumonia in Hubei Province, China

**DOI:** 10.1101/2020.04.01.20050526

**Authors:** Jianfeng Li, Linyuan Zhang, Zhihua Ren, Caihong Xing, Peihuan Qiao, Bing Chang

**Affiliations:** Beijing Municipal Institute of Labour Protections, Beijing Academy of Science and Technology. Central Road, North 4th Ring Road, Haidian District, Beijing, 100083, China; National Institute of Occupational Health and Poison Control, Chinese Center for Disease Control and Prevention. No. 29 Nanwei Road, Xicheng District, Beijing, 100050, China; National Meteorological Information Center, China Meteorological Administration Meteorological Data Center. No.46 Zhongguancun South Avenue, Haidian District, Beijing 100081, China

**Author notes:** Corresponding author: Bing Chang Ph.D., E-mail addresses. Jianfeng Li and Linyuan Zhang should be considered joint first author.

**Keywords:** 2019-nCoV, COVID-19, meteorological factors, pandemic, average water vapor pressure, average minimum temperature, average atmospheric pressure

## Abstract

**Objective:** many potential factors contribute to the outbreak of COVID-19.It aims to explore the effects of various meteorological factors on the incidence of COVID-19.

**Methods:** Taking Hubei province of China as an example, where COVID-19 was first reported and there were the most cases, we collected 53 days of confirmed cases (total 67773 cases) and ten meteorological parameters up to March 10. Correlation analysis and linear regression were used to judge the relationship of meteorological factors and increment of COVID-19 confirmed cases.

**Results:** Under 95% CI, the increment of confirmed cases in Hubei were correlated with four meteorological parameters of average pressure, average temperature, minimum temperature and average water vapor pressure (equivalent to absolute humidity).The average pressure was positively correlated with the increment (r=+0.358).The negative correlations included average temperature (r=-0.306), minimum temperature (r=-0.347), and average water vapor pressure (r=-0.326). The linear regression results show if minimum temperature increases by 1□, the incremental confirmed cases in Hubei decreases by 72.470 units on average.

**Conclusion:** Statistically, the incidence of COVID-19 was correlated with average pressure, average temperature, minimum temperature and average water vapor pressure. It is positively correlated with the average pressure and negatively correlated with the other three parameters. Compared with relative humidity, 2019-nCov is more sensitive to water vapor pressure. The reason why the epidemic situation in Hubei expanded rapidly is significantly related to the climate characteristics of low temperature and dryness of Hubei in winter.

## 1 Introduction

On December 2019, 27 cases of unknown pneumonia broke out in Wuhan, Hubei province (Wuhan City Health Committee, 2019). On January 9, 2020, Chinese CDC reported to the world that a new coronavirus (2019-nCoV) was detected in these serious cases (Corman et al., 2020). On January 22, 2019, Hubei Health Commission announced the outbreak of new coronavirus pneumonia (COVID-19) to the public and shared all data on disease transmission (Holmes, 2020). On March 11, when the new coronavirus disease (COVID-19) spread to 114 countries and the confirmed cases around the world were close to 120,000, WHO declared the disease to be pandemic (WHO, 2020).

COVID-19 is caused by severe acute respiratory syndrome coronavirus type 2 (SARS-CoV-2/2019-nCoV), SARS-CoV-2 belonging to β coronavirus, is enveloped virus with positive-strand RNA genome (Chan et al.,2020). 2019-nCoV infection include fever, cough, myalgia or fatigue, dyspnea and bilateral ground glass shadow of chest CT scan, and few obvious upper respiratory symptoms (such as snot, sneezing, or sore throat), it indicated that the virus mainly infected the lower respiratory tract (Chan et al.2020;Huang et al.,2020). It is worth noting that severe and critically ill patients may not have obvious fever, and may have acute respiratory distress syndrome, septic shock, metabolic acidosis, hemorrhage and Coagulation dysfunction, even death (Huang et al.,2020). Men over 80 years old and patients with complications are associated with an increased risk of death, and the characteristics of the dead patients are very consistent with MuLBSTA (Chen et al.,2020). The WHO’s research and development blueprint and the global organization for infectious disease prevention and control research cooperation (GLOPID-R) have determined to give priority to the study of virus pathogenesis and clinical characteristics, prevention and control of infection and plans for candidate therapies (GloPID-R,2020).

Respiratory virus infection can occur in the following ways:(a) direct/indirect contact, (b) droplet spray in short-distance transmission, or (c) long-distance transmission by aerosol, i.e. air transmission (Brankston et al.,2007). Research shows that COVID-19 is transmitted through direct contact or droplets (Chan et al.,2020; Li et al., 2020;Wang et al.2020) while air transmission is not excluded (van Doremalen et al., 2020). Environmental factors affect host susceptibility by regulating respiratory tract defense mechanism, and affect the viability and transmission of respiratory tract viruses in the environment through aerodynamic mechanism. Human behavior will affect the contact rate between infected individuals and susceptible individuals. Both epidemiology and laboratory studies show that meteorological indicators are important factors for the transmission and survival of respiratory tract infection viruses (enveloped and non-enveloped viruses) (Chan et al. 2011; Ma et al.,2020;Shaman & Kohn, 2009; Xie & Zhu.,2020;Yao et al.,2020;Yuan et al.,2006;Yusuf et al.,2007). In particular, it is unclear that the meteorological factors how to influence on the activity of 2019-nCoV, and then change the spread of the virus through a specific process, and to affect the morbidity. However, we believe that the relationship between meteorological factors and the characteristics of COVID-19, as well as the mechanism of meteorological factors on the transmission route of 2019-nCoV, are of practical significance to prevent and control the epidemic disease, especially the nosocomial infection and community transmission, and to predict the extinction of the virus.

Due to the different characteristics of virus transmission in different latitudes, Hubei province is used instead of China or Wuhan city with appropriate spatial resolution (that is, at the provincial level rather than at the national level, nor at the municipal level), and analysis of observation data is of much concern for understanding the global durability of viruses and formulating appropriate prevention and control measures. This study designed the correlation analysis between the daily confirmed cases increment of COVID-19 in Hubei province and 10 meteorological parameters and the regression analysis of four relevant parameters, combined with the meteorological characteristics of Hubei province, in order to find out the associative clues of meteorological factors on the occurrence, development and extinction of COVID-19 epidemic.

## 2 Methods

### Ethics statement

COVID-19 surveillance data used in this study was collected from the official website of Health Commission of Hubei Province. Hubei Meteorological Observatory data were obtained from the Meteorological Data Center of China Meteorological Administration. Therefore, ethical approval was not required.

### 2.1 Date collection

#### (1) Meteorological Observatory data

Meteorological parameters are from data collected from representative meteorological stations in Hubei province provided by the meteorological data center of China meteorological administration. The available data period is from January 1, 2020 to March 10, 2020 (100 daily meteorological data). Meteorological observatory data include: average air pressure (hPa), average temperature (°C), maximum temperature (°C), minimum temperature (°C), average water vapor pressure(WVP) (hPa) (equivalent to absolute humidity(AH)), average relative humidity(RH, %), average wind speed (m/s), precipitation (mm), total solar radiation (0.01 0.01 MJ / m^2^), maximum solar irradiance (W/m^2^). Afterwards, it uses Ave_Pre, Ave_Temp, Max_Temp, Min_Temp, Ave_WVP, Ave_RH, Ave_WS, Prep, Tot_SR and Max_SI to refer to them respectively.

#### (2) Number of confirmed cases

Case data include Hubei’s cumulative confirmed cases, Hubei’s cumulative death, and Hubei’s incremental cases,namely HB_CumN, HB_CumD, and HB_DeltaN respectively, were collected from the official website of Wuhan Health Committee. This study uses January 17, 2020 as the starting time to retrieve data, as of March 10, 2020, for the reason that COVID-19 had been clearly diagnosed and the great improvement of detection ability and speed. In addition, 1) on February 12, 2020, the number of newly incremental cases in Hubei province surged explosively to 14840 (including 13332 clinically diagnosed cases), in which 13436 cases were in Wuhan. 2) The incremental confirmed cases in January 19, 2020 were 0. Here, the above two days are outliers and are eliminated in correlation analysis and regression analysis.

### 2.2 Selection of statistical methods

#### (1) Correlation analysis

Spearman rank correlation coefficient was used in the correlation analysis for binary variables. The calculation formula is as follows:

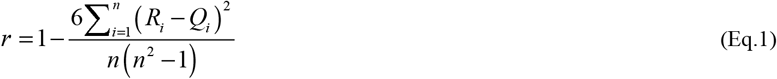

In this research, it sets confidence level P = 0.05, CI = 95%. SPSS 15.0 (No. c23f0922c168a5262283)was used to statistical analysis.

#### (2) Linear regression

On the basis of correlation analysis, we estimate the parameters in the regression using the Ordinary Least Square (OLS) method. Incremental cases and four meteorological parameters are further selected for unary linear regression.

The prediction model of linear regression is:

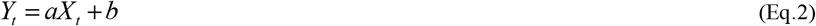

In the formula,

*X*_*t*_ represents the period *t* independent variable;

*Y*_*t*_ represents the dependent variable of period *t*;

a and b represent the parameters of a linear regression equation. Obtained by the following formula:

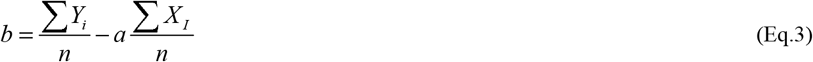

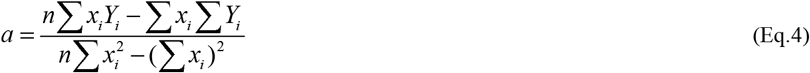

For the regression analysis of quantitative data, four prerequisites must be met: the linear relationship between incremental cases (dependent variables) and certain meteorological parameters, and each parameter is independent of each other, the residual of incremental cases satisfies the normality, and the residual of incremental cases satisfies the homogeneity of variance. If linear regression results cannot be obtained, generalized linear regression is considered.

## 3 Results

### 3.1 Basic statistics of incremental cases and meteorological parameters

Table 1 summarizes the basic statistical results of incremental cases and meteorological parameters. As of March 10, 2020, there are 67773 confirmed cases in Hubei province. The minimum number of incremental cases was 13 and the maximum was 4823, which occurred on February 13, 2020. From January 1, 2020 to March 10, the average air pressure range is 1006.0-1031.2 hPa, the average is 1021.7hPa, (SD.5.1 hPa), and the fluctuation range is small. The average wind speed ranges from 0 to 5.0 m/s and the fluctuation range is small too. The maximum rainfall is 38.9mm, which occurs in January 9, 2020 and has no obvious regularity. The average temperature, the maximum temperature and the minimum temperature are 6.9°C, 11.6°C and 3.4°C respectively, the standard deviation of the maximum temperature is 5.0°C > that of the minimum temperature 4.3°C > that of the average temperature 4.0°C. The average WVP and the average RH are both related to the water loading in the air, and the mean ± standard deviations are 8.3 ± 2.4 (hPa) and 83.0 ± 8.2 (%), respectively. Ultraviolet intensity is not the routine monitoring items in the meteorological station. Therefore, the daily total solar radiation and the maximum solar irradiance are selected as two auxiliary parameters, and their mean values are 757.7± 587.3 (0.01 MJ/m^2^) and 456.6±274.2 (W/m^2^), the standard deviations are (0.01 MJ/m^2^) and 274.2 (W/m^2^), respectively. Obviously, the volatility of the values is very strong.

**Table 1.**
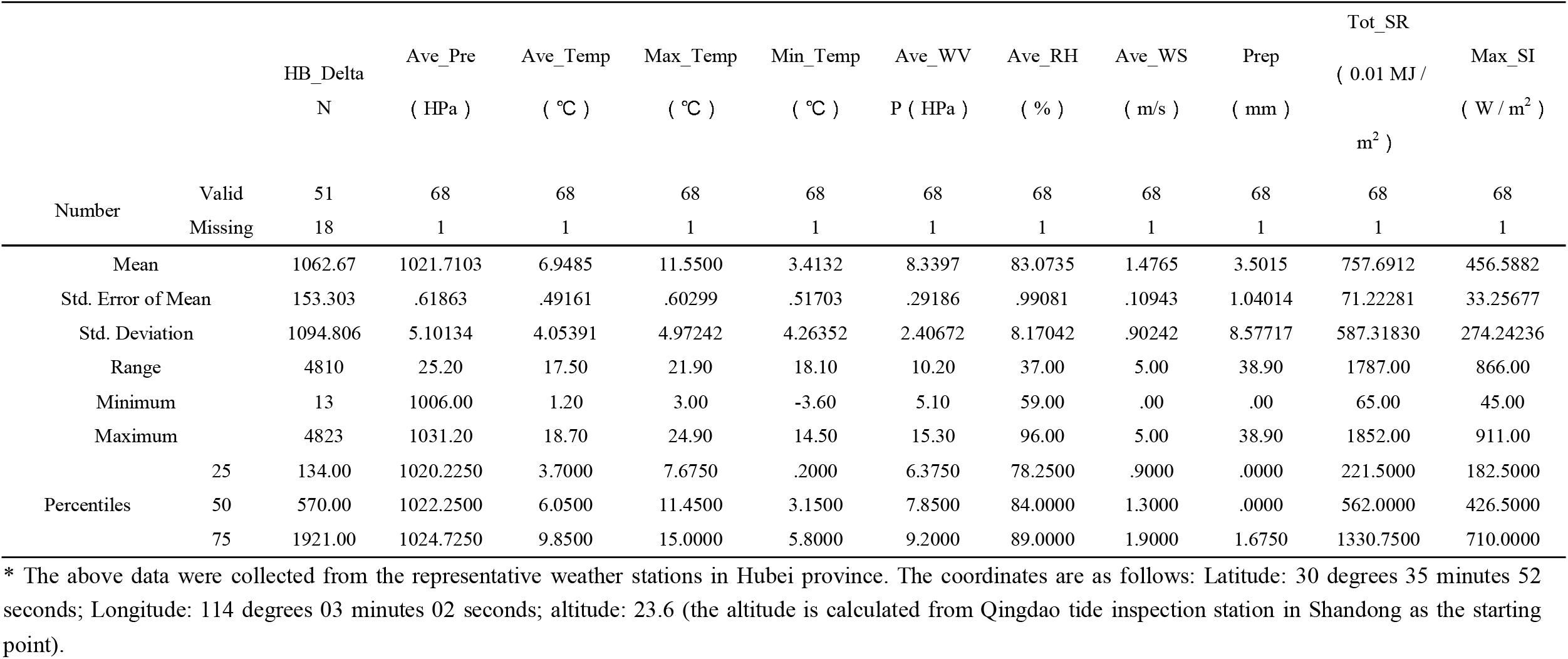
Basic statistical analysis of HuBei delta number and weather parameters

Figure 1 shows the frequency statistics of incremental cases and meteorological parameters, among which the normality of average air pressure, average temperature, minimum temperature, average RH and average wind speed is better. The total solar radiation and the maximum solar irradiance present a saddle-shaped bimodal structure.

**Figure 1.**
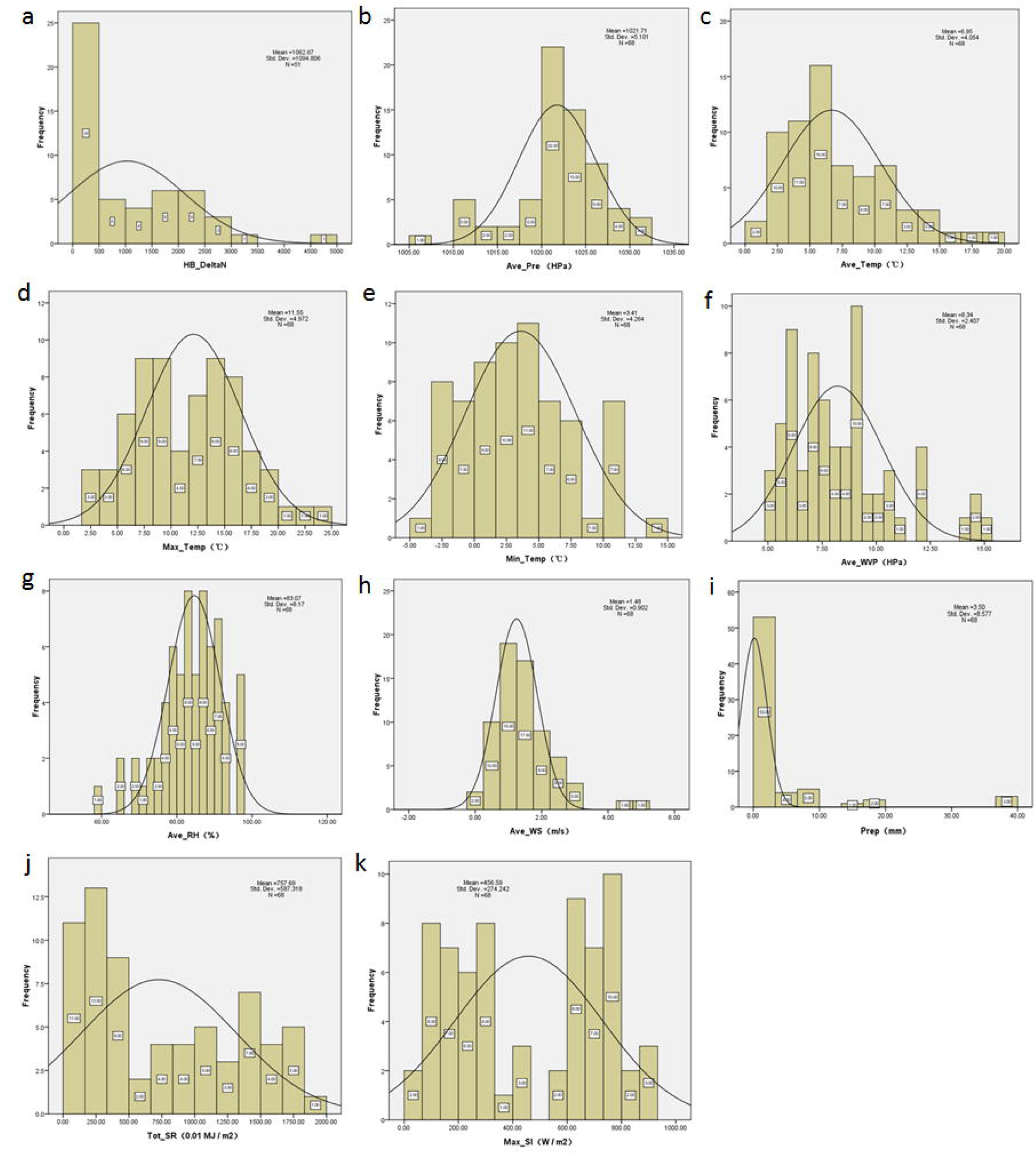
includes 11 sub-figures. (a)demonstrates the frequency statistics of incremental cases in Hubei province; (b)-(k) demonstrates the frequency statistics of average pressure, average temperature, maximum temperature, minimum temperature, average water vapour pressure, average relative humidity, average wind speed, precipitation, total solar radiation, maximum solar irradiance

Figure 2 (a) shows the tendency of incremental cases changing with time. Obviously, it is a single-peak structure. Figure 2(b-d) shows the tendency of average temperature, average WVP and the maximum solar irradiance with time. Figure 2(b) as for average temperature-time pair, it can be seen that there is a fluctuating process after about January 10, 2020. Figure 2(c) as for average WVP-time pair, after January 10, 2020, also has a fluctuating process, but the trend is relatively gentle. Figure 2(d), there are two parallel tracks for the maximum solar irradiance-time pair, the general trend is rising but gentle.

**Figure 2.**
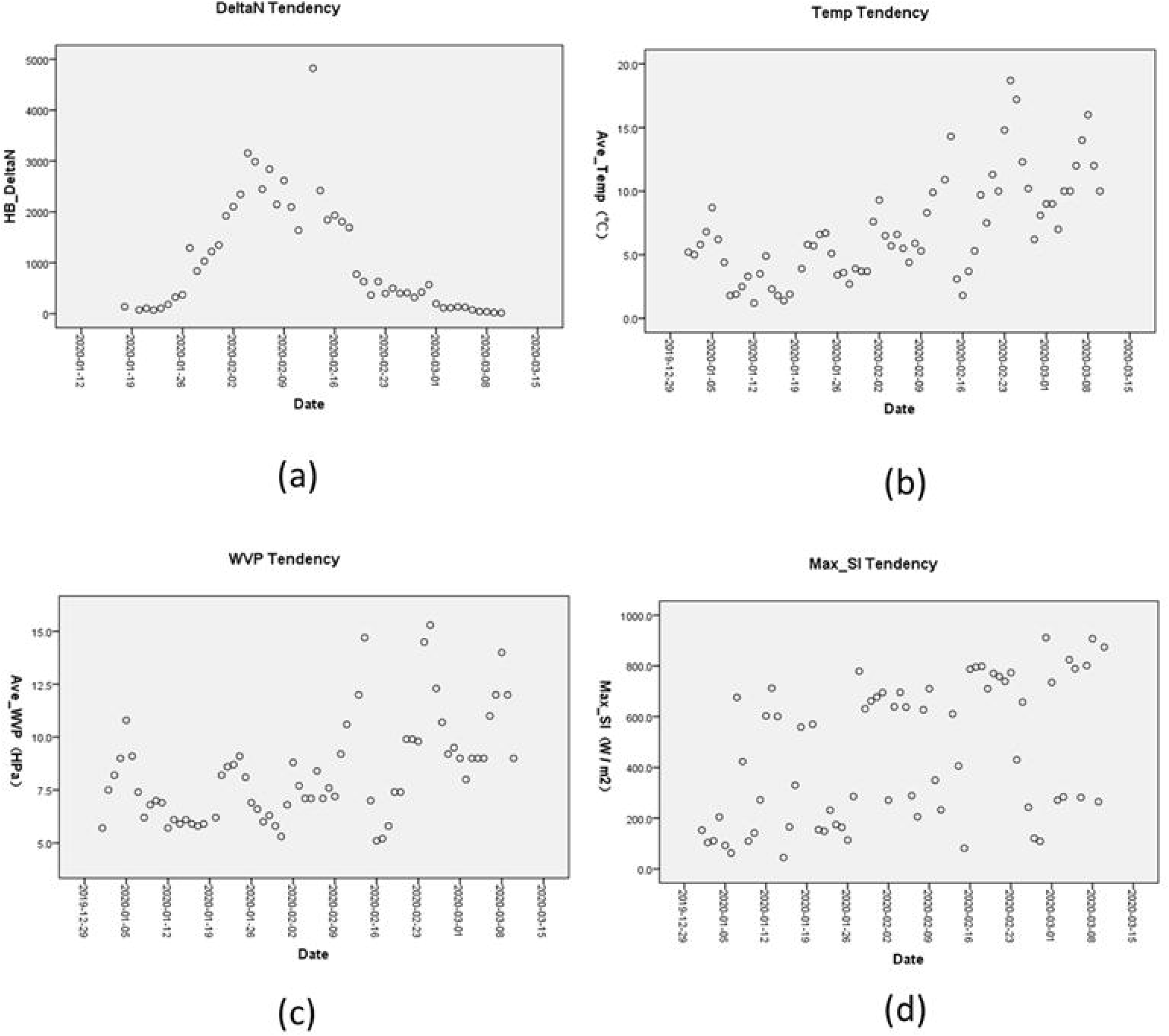
includes 4 sub-figures. (a) to (d) shows the tendency of incremental cases, average temperature, average water pressure and the maximum solar irradiance changing with time

### 3.2 Correlation Analysis between incremental cases and meteorological parameters

Incremental cases in Hubei province is correlated with four meteorological parameters including average air pressure, average temperature, minimum temperature and average WVP (P<0.05). Among them, there is a positive correlation with the average air pressure with the correlation coefficient r = + 0.358. Negative correlation include average temperature, minimum temperature, average WVP with its coefficient r =-0.306,-0.347, −0.326, respectively (Table 2, Figure 3). From this, it can be judged that the increment of cases will be inhibited by the increase of temperature and water loading in the air. At the same time, the increase of average air pressure may increase the reproduction rate of the virus.

**Table 2.**
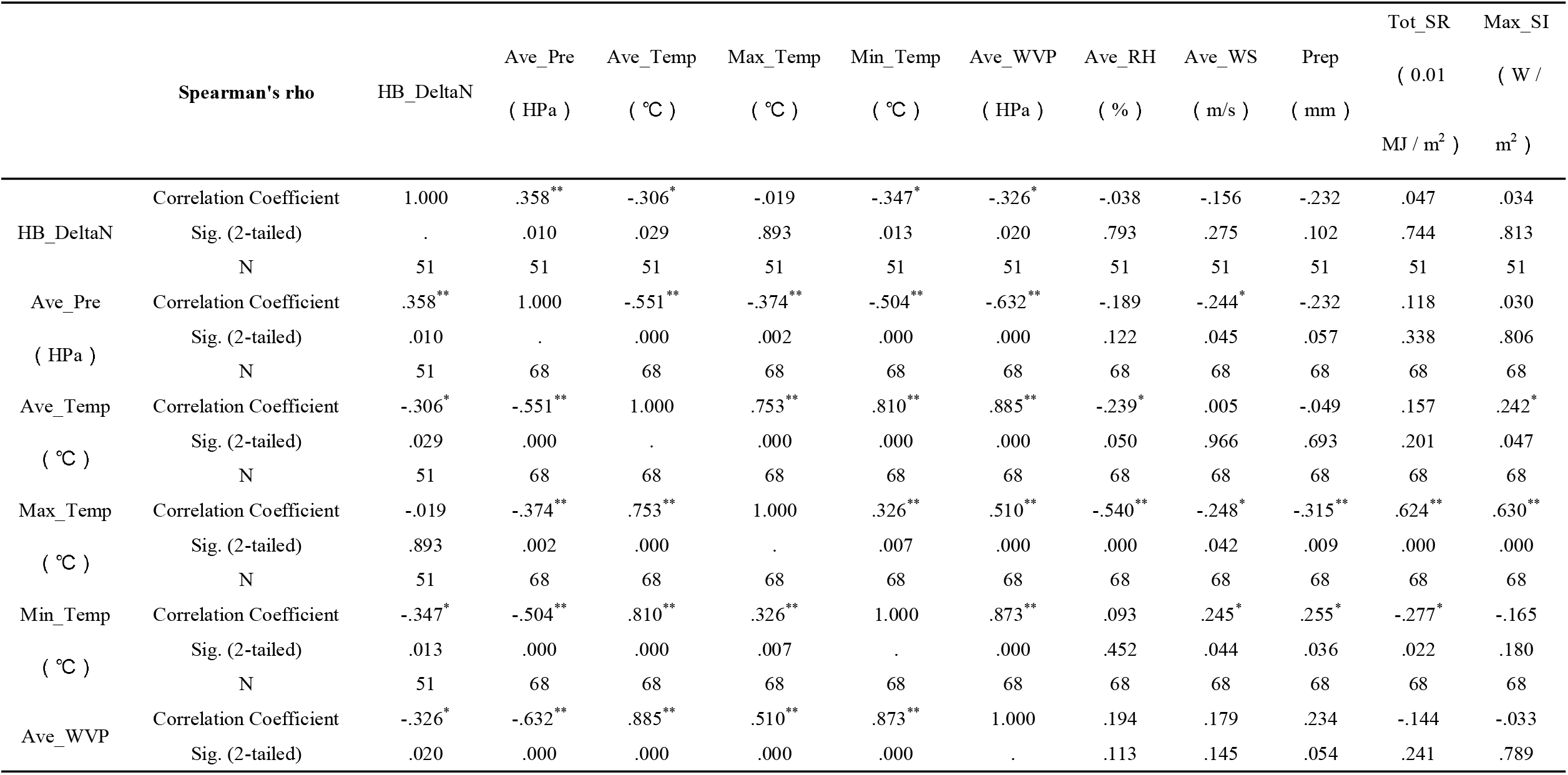
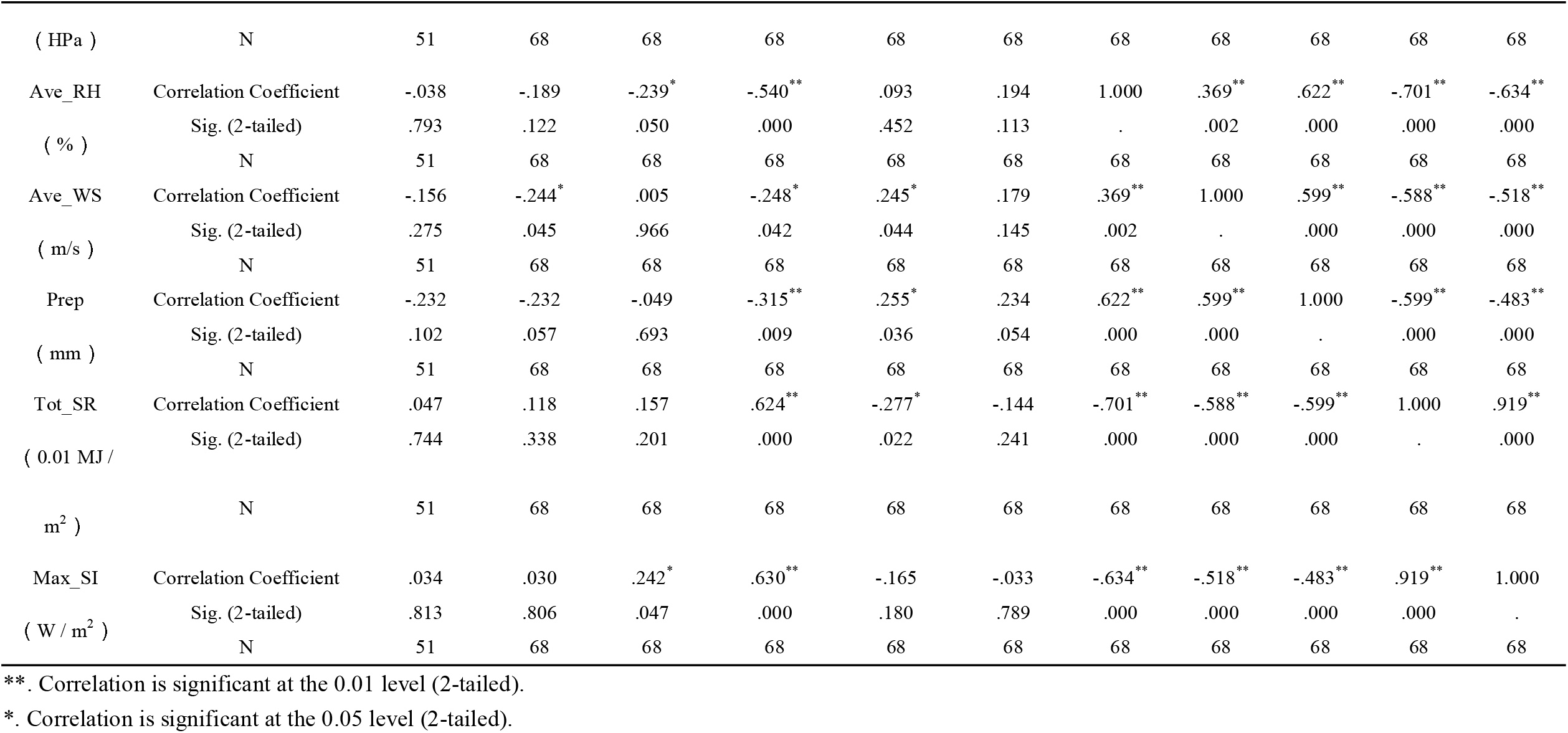
Cross-Relationship between Hubei delta number with weather parameters

**Figure 3.**
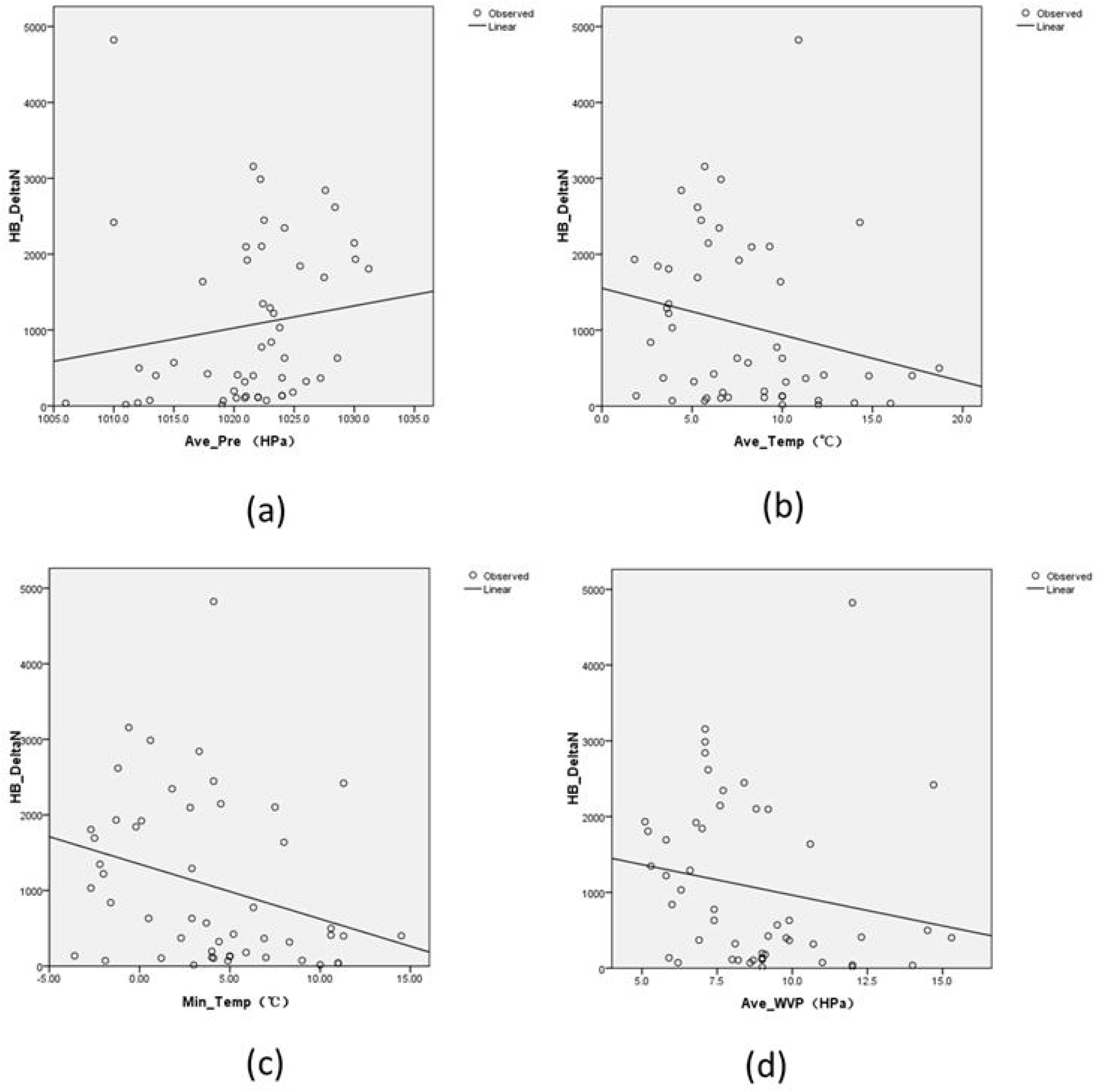
includes 4 sub-figures. (a) to (d) shows curve fitting results of HB_DeltaN with average pressure, average temperature, minimum temperature and average water vapour pressure

### 3.3 Regression analysis of incremental cases and meteorological parameters

Through parameter selection in the way of entering in sequence, finally only the minimum temperature and incremental cases can establish a statistically significant linear regression model (95%CI, F=4.835>F_n-m-1_=4.043,P<0.05), and the equation is as follows:

HB_DeltaN = 1348.425-72.470 × Min_Temp

Thus, if Min_Temp increases by 1°C, HB_DeltaN will decrease by 72.470 units on average. In other words, the result shows that the minimum temperature changes by 1 degree, and 72 incremental cases will change in the opposite direction. Let HB_DeltaN = 0, and get Δmin_temp = 18.6°C, therefore, minimum temperature rises 18.6°C on the basis of the mean value of 3.4°C, that is, if the average minimum temperature reaches 22°C, case increment statistically shrink to zero (Table 3). In addition, the residual analysis of the regression model shows that the residual conform to the normal distribution. The distribution of points in scatter plot, in which the X axis is ZRESID(regression starndardized residual), the Y axis is ZPRED(regression standardized predicted value), is uniform, indicating that the variances are homogeneous (Figure 4).

**Table 3.** Linear Regression Model Summary and Parameter Estimates

| Dependent Variable | Explanatory variable | Parameter Estimates |  | Equation Expression | ANOVA | R2 | P |
| --- | --- | --- | --- | --- | --- | --- | --- |
| | | Constant | b | | $F > F_{n-m-1} = 4.043$ | | $\alpha = 0.05$ |
| HB_DeltaN | Min_Temp | 1348.425 | -72.470 | $HB\_DeltaN = 1348.425 - 72.470 \times Min\_Temp$ | 4.835 | 0.090 | 0.033 |
| HB_DeltaN | Ave_Pre | -28797.5 | 29.239 | × | 1.117 | 0.022 | 0.296 |
| HB_DeltaN | Ave_Temp | 1552.099 | -61.511 | × | 2.668 | 0.052 | 0.109 |
| HB_DeltaN | Ave_WVP | 1771.235 | -80.843 | × | 1.755 | 0.035 | 0.191 |

**Figure 4a.**
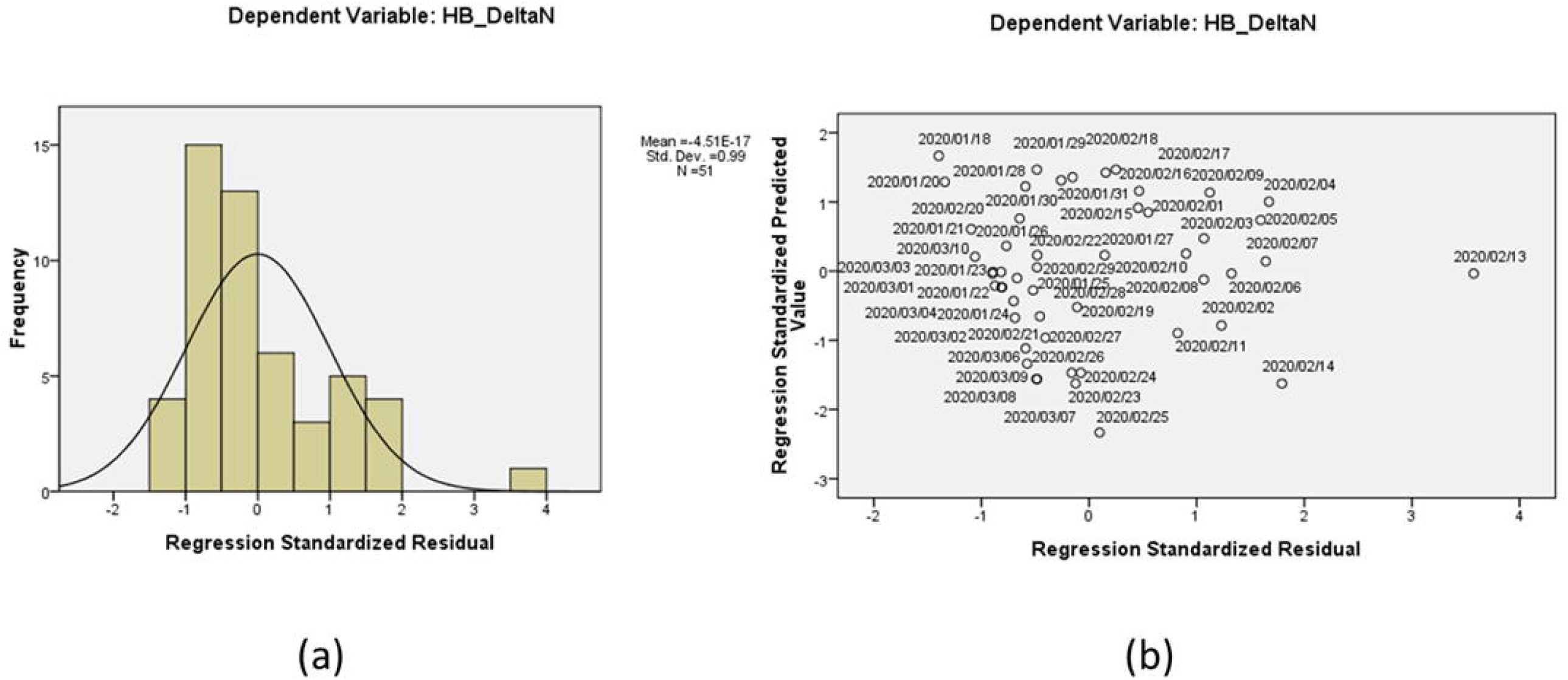
shows the normality of the residual of incremental cases satisfies. Figure 4b the homogeneity of variance for the residual of incremental cases

## 4 Discussions

Global attentions focus the potential impacts on the COVID-19 evolution process and fate exerted by seasonal variation in meteorological parameters. The outbreak of COVID-19 in Hubei province from the end of 2019 to the beginning of 2020 has obvious correlation to meteorological factors. By using the Spearman rank correlation coefficient, it discussed the correlation between daily incremental cases and meteorological factors in Hubei province. The results showed that the average temperature, the minimum temperature, WVP and air pressure were significantly correlated with the daily incremental cases. Incremental cases is positively correlated with atmospheric pressure and negatively correlated with average temperature, minimum temperature and WVP. Further, a linear regression model disclosed the linear relationship between the cases and the minimum temperature.

It found that the average temperature and the minimum temperature were negatively correlated with the COVID-19 incremental cases in Hubei province. The unary linear regression results showed that the minimum temperature increased by 1°C and the incremental cases decreased by 72 cases. This is consistent with studies of other respiratory viral infectious diseases. Studies have reported that SARS-CoV is sensitive to temperature and relatively stable at low temperatures. Higher relative humidity (RH)(>95%) and high temperature (e.g. 38°C) has synergistic effect on the inactivation of SARS-CoV activity, while lower temperature and lower humidity can prolong the survival time of virus on contaminated surface (Chan et al.,2011). The number of daily SARS cases is negatively correlated with the highest and/or lowest temperature (Bi et al.,2007), and the rate of SARS recurrence is also negatively correlated with the daily average temperature (Cai et al.,2007). During the epidemic period, in the days with lower temperature, the risk of increasing the daily incidence of SARS is 18.18 times higher than that in the days with higher temperature (Lin et al.,2006). Regression analysis showed that there was a significant correlation between the average temperature (r =-0.45, P<0.001) recorded 13 to 17 days before SARS was confirmed and the number of clinically diagnosed cases (Yuan et al.,2006).

In addition to SARS, meteorological factors affect seasonal influenza prevalence. Studies have found that low temperature may prolong the survival time of influenza virus (Cheng et al., 2016) and may lead to more influenza transmission (Yu et al., 2013). For every 1°C drop, the risk of influenza infection in Finland increases by 11% (Jaakkola et al.,2014). In addition, studies revealed that a 1°C drop in temperature in Hong Kong leads to an 8.55% rise in influenza cases and a 32.14% rise in UK (Wang et al., 2017).

Recently studies demonstrated that relationship between temperature and COVID-19 mortality. Every increase of 1°C in the diurnal temperature range caused COVID-19 mortality 2.92% increase (Ma et al.,2020). Another study reported that the relationship between the average temperature and COVID-19 confirmed cases was approximately linear in the range of <3°C, while it tended to be flat above 3°C. When the average temperature is lower than 3°C, the number of confirmed cases per day increases by 4.861% (95% CI:3.209% ∼6.513%) (Xie & Zhu, 2020). This result is contrary to our result. This study used daily confirmed cases and meteorological data (37 days in total) from multiple onset areas (122 cities) from January 23 to February 29, 2020. The difference is that two-month interval of observation included 53 days effective data, and we only use the data of Hubei province (including Wuhan, Xiaogan, Huanggang, and other a total of 17 cities or regions), covering a total of 67773 confirmed cases. The explanation of the different results still needs to be discussed in depth. There is also a recent report on the relationship between COVID-19 and meteorological factors in 224 cities in China, among which the cumulative incidence rate and temperature have not found a significant correlation (Yao et al.,2020). We believe that meteorological factors including temperature may be more relevant to the daily increasing case number rather than the cumulative number. In addition, the difference between the total number and the onset period in each city may also be a related factor affecting the results.

We found that the daily increasing case number of COVID-19 cases was associated with WVP rather than RH. The correlation between the daily increasing case number and RH is not statistically significant. Our study conducted curve fitting between average WVP and delta number (daily case increment) using S-curve. The S-curve model HB_DeltaN = EXP(4.235+15.919/ Ave_WVP) was obtained (95%CI, F = 6.927> F_n-m-1_ = 4.043,P<0.05). It could be seen from the regression equation that when the explanatory variable (average WVP) increased, the exponential part of the equation decreased, thus the response variable (case increment) decreased, this further illustrated that there is a negative correlation between the average WVP and the case increment, and goodness of fit R^2^ = 0.124(P<0.05), which indicated that WVP can explain the change of 12.4% cases. Our results proved that compared with RH, WVP is more sensitive to COVID-19 occurring in epidemic areas of Hubei province. At the same time, this result proved the theory of meteorological factors correlation with viral respiratory infectious diseases proposed during the onset of influenza (Shaman & Kohn,2009;Shaman et al.,2010) and introduced new evidence in COVID-19 viral transmission.

Shaman & Kohn (2009) re-analyzed the correlation between the survival and transmission of respiratory viruses and WVP (equal to AH), rather than RH. They used the data from previous animal influenza transmission experiments, the RH is converted into WVP by using Clausius–Clapeyron relation, and it is found that the more strong correlation between WVP and influenza transmission and influenza virus survival. Shaman & Kohn deemed that, for influenza virus transmission and survival activity, explanatory ability of WVP was 50% and 90% respectively, for RH, it was 12% and 36% (Shaman & Kohn, 2009). In the research on mortality model related to influenza, it was found that the change of WVP was the root cause of seasonal changes in influenza, while RH was not (Shaman et al.,2010). Since then, the theoretical hypothesis of respiratory virus-induced diseases with WVP as the main regulatory factor has been established. After that, causal analysis of global influenza incidence data shows that temperature change and low AH are more likely to lead to changes in influenza activity than actual low temperature. According to their research, the conclusion is that WVP is a stronger driving factor than RH (Deyle et al.,2016).

China’s epidemiological studies proved that influenza virus is negatively correlated with temperature and WVP (Sun et al., 2018). In sub-frigid climate (Norway), WVP below 4 hPa on average may trigger influenza (Kolberg et al., 2019). Studies in tropical/subtropical regions of China have found that, WVP was negatively correlated with the incidence ofH1N1pdm09 (Influenza A) and Yamagata (Influenza B) (Pan et al., 2019). The study of Kormuth et al. (2018) showed that the infectivity to influenza virus in droplets and aerosols had no difference under different RH, that was to say, it had nothing to do with RH (Kormuth et al.,2018). This undoubtedly brought new challenges to RH and respiratory virus survival and transmission. The relevant mechanism of WVP may be due to the degeneration of virus lipoprotein. Higher level of WVP will lead to surface inactivation of lipid-containing virus (Yang et al.,2012), it is related to the phase transition of phospholipid bilayer membrane, which causes the peptide to cross-link at the air-water interface. However, there is still a lack of comprehensive understanding of the mechanism of WVP affecting virus survival (Peci et al.,2019).

In previous studies of SARS, the number of SARS cases in Beijing was significantly negatively correlated with RH (r= −0.784,P<0.05). Regression analysis showed that there was a significant correlation between the RH recorded 13 to 17 days before SARS was confirmed and the number of clinically diagnosed cases (Yuan et al.,2006). Others found that RH did not appear to play a role in the transmission of SARS-CoV (Cai et al.,2007). No relevant reports of WVP were found in previous studies of SARS. The latest research in COVID-19 found that in Wuhan, the WVP (equal to AH) is related to the COVID-19 mortality rate decrease. The WVP increases by 1 unit, the maximum death decline was-11.41% (95% CI:-19.68%∼-2.29%)(Ma et al.2020). However, no research on the effect of WVP on the number of confirmed cases has been reported so far. At the same time, no positive result involving the influence of RH on the number of confirmed cases has been found (Yao et al.,2020;Xie & Zhu et al.,2020).

We found a positive correlation between incremental cases of COVID-19 and atmospheric pressure. The average atmospheric pressure in Hubei province is 1021.7(1006.0∼1031.2)hPa, and correlation coefficient r=+0.358 (P<0.05). Previous studies believed that atmospheric pressure was positively correlated with influenza virus (Soebiyanto et al., 2010). Atmospheric pressure may be the cause of influenza (Sundell et al.,2016). RSV activity increased with the increase of atmospheric pressure (Hervás et al., 2012). In the United States, a study analyzing the relationship between meteorological factors and RSV activity in 9 cities under different climate types showed that in Delhi region, air pressure was the main relevant factor of RSV total amount, which was associated with 22% RSV activity (Yusuf et al.,2007). In Beijing (Yuan et al.,2006) and Hong Kong (Bi et al.,2007), atmospheric pressure was positively correlated with the spread of SARS during the SARS epidemic. Atmospheric pressure is the air weight over unit area, i.e. air density. When the human body expels respiratory viruses, the increase of atmospheric pressure may lead to the decrease of the diffusion range of liquid drops and the increase of the number of viruses per unit area. As the infectivity of viruses depends on the number of viruses per unit area, if the concentration per unit area of the virus is insufficient due to the increase of atmospheric pressure in the transmission of RSV, infection will not occur. Similarly, high diffusion may produce low concentrations of infectious particles (Collins & Kennedy,1999). Therefore, a balance must be maintained between virus concentration and its dispersion to achieve effective infectivity (Roy & Milton,2004), but the detailed mechanism needs further study. However, for COVID-19, no research results have been found to explore the relationship between atmospheric pressure and confirmed cases.

We found for the first time the correlation between the minimum temperature, WVP and new cases of COVID-19. According to the analysis of its relevant mechanism, non-enveloped viruses (HRV and HEV) that cause respiratory tract infection may spread through close contact or droplet, rather than mainly through aerosol, therefore, the transmission and survival of the virus are relatively less dependent on meteorological factors (Sundell et al.,2016). However, enveloped virus (influenza, RSV, SARS-CoV, 2019-nCoV) has greater dependence on meteorological factors. At the same time, previous studies have shown that envelope protein (E) of coronavirus causing severe acute respiratory syndrome is directly related to virus virulence (Schoeman & Fielding.2019). We believe that envelope proteins that may be closely related to 2019-nCov pathogenicity are sensitive to minimum temperature and WVP, and they are more vulnerable to climatic factors, especially temperature (including average temperature and minimum temperature) and WVP. This inference is of theoretical significance for understanding meteorological factors and viral respiratory infectious diseases. Of course, more evidence is needed to support it in the future.

During the COVID-19 epidemic, human-to-human transmission was still in a phase dominated by community and hospital infections. At this time, meteorological factors, especially low temperature and low WVP had related effects on the *in vitro* viability, the quantity per unit area and transmission distance of 2019-nCov. We believe that meteorological factors influence on virus viability and the contribution capacity of changing the transmission is 9%(minimum temperature) and 12.4%(WVP), respectively. These do not belong to manual measure to control transmission such as virus elimination, washing hands, wearing masks, and keeping social distance. Meteorological factors play a role in the spread of the virus from patients (including asymptomatic infected people) to unprotected people (including susceptible and non-susceptible people). It will involve biochemistry, photochemistry and aerodynamics.

Considering the statistical period of 3 months, the actual effective data is only 53. In the case of limited data, there are 4 parameters with obvious correlation, which is completely reasonable to believe that, first, with the increase of effective observations, the correlation between parameters will gradually appear; secondly, case increment is very sensitive to meteorological parameters, and analytical method using meteorological parameters is effective and trustable; thirdly, in addition to the minimum temperature, the other three parameters failed to establish a linear regression model. Since different meteorological factors have different mechanisms for virus survival and transmission, each index has its own characteristics, so it is necessary to establish different regression equation fitting analysis of their characteristics, and to further study the multi-parameter coupling mechanism.

This study has some limitations. First, we did not distinguish the gender, age, health status and other information of the patients in the case, nor did we consider the host factors that play a role in the spread of the disease, such as immunity, this may result in bias of the derived results. In addition, we cannot rule out the possible influence of some mixed factors such as human movement and air pollution. Second, previous studies have shown that viruses that cause respiratory infectious diseases are sensitive to climate. Climate factors may affect the survival and transmission of viruses in the environment, host susceptibility and exposure possibilities. We found that meteorological factors played an important role during the COVID-19 in Hubei province, but we are also very clear that, as scholars have known, meteorological parameters can only explain no more than 30% of influenza activity changes (Monamele et al.,2017), and there are still many problems to be confirmed about 2019-nCoV. Third, the impact of social distancing measures on the epidemic situation should be considered in the study. This measure is an important part of the public health response to COVID-19. The corresponding measures taken in Hubei province and Wuhan City have seen actual results.

## 5 Conclusions

First of all, in January of each year, the overall temperature and humidity in Hubei province are at the lowest level throughout the year, and it is in this month that COVID-19 erupted on a large scale. The time range is from January 1, 2020 to March 10, with 67773 confirmed cases cumulatively. Meteorological factors (parameters) provide evidence related to the incidence of COVID-19. Correlation analysis showed that average atmospheric pressure, average temperature, minimum temperature and average WVP were significantly correlated with daily increase in COVID-19 cases. Obviously, different climate factor indicators may affect the occurrence of COVID-19 epidemic. Although this conclusion comes from the evolution of confirmed cases in COVID-19 in Hubei province of China, they still show that, various results are compatible within the current knowledge range of COVID-19 epidemics. The significance of this work is that increasing the decline of cases may be affected by changes in meteorological factors that limit the ability of virus transmission. The differences in meteorological factors should be taken into account when predicting the epidemic scale during the global transmission of 2019-nCoV and formulating prevention and control strategies.

Secondly, we analyzed the possible relationship between meteorological factors and COVID-19 attacks from epidemiological statistics, as well as the influence of meteorological parameters on the transmission and survival of viruses causing respiratory diseases, and analyzed its possible mechanism. We demonstrated the epidemiological correlation between WVP and COVID-19. At the same time, we speculate that compared with non-enveloped virus, enveloped virus” 2019-nCoV” is more sensitive to climatic factors, especially temperature (including average temperature and minimum temperature).

Third, meteorological factors may be one of the influencing factors causing the 2019-nCoV epidemic. The study used a linear regression model to explain the relationship between incremental cases and the lowest temperature. The results show that for Hubei province (epidemic area), for every 1°C increase in the lowest temperature, 72 incremental cases are reduced, and when the lowest temperature reaches 22°C, the increased cases are significantly reduced. Therefore, it is speculated that in other epidemic areas in the world, if the minimum temperature rises above a certain threshold, the number of new cases may drop dramatically.

## Data Availability

The data produced in this study can be found in the Supplementary information.

## Acknowledgements

No one to thank in particular

**The authors have no conflicts of interest**.

## Supporting information

Supplementary information contains an Excel file with datasheet containing the meteorological observatory data and number of confirmed cases from January 1, 2020 to March 10, 2020 in Hubei province of China.

## Data accessibility

The data produced in this study can be found in the Supplementary information. The datafile includes an Excel file and a Word file which describes the data.

## Supporting information-data description

### (1) Meteorological Observatory data

The available data period provided from the Meteorological Data Center of China Meteorological Administration is from December 1, 2019 to March 10, 2020 (100 daily meteorological data). The actual data selected starts on January 1, 2020 and ends on March 10, 2020. Meteorological observatory data include: average air pressure (hPa), average temperature (°C), maximum temperature (°C), minimum temperature (°C), average water vapor pressure (hPa) (equivalent to absolute humidity), average relative humidity (%), average wind speed (m/s), precipitation (mm), total solar radiation (0.01 megajoules/square meter), maximum solar irradiance (Watt/square meter). Afterwards, it uses Ave_Pre, Ave_Temp, Max_Temp, Min_Temp, Ave_WVP, Ave_RH, Ave_WS, Prep, Tot_SR and Max_SI to refer to them respectively.

### (2) Number of confirmed cases

This study uses January 17, 2020 as the starting point of time to retrieve data. Case data include Hubei’s cumulative confirmed cases, Hubei’s cumulative death, and Hubei’s incremental cases, namely HB_CumN, HB_CumD, and HB_DeltaN respectively. February 12, 2020 and January 19, 2020 are outliers.

